# Vestibular Function Predicts Prefrontal and Sensorimotor Cortical Gray Matter Volumes in a Cross-Sectional Study of Healthy, Older Adults

**DOI:** 10.1101/2022.11.20.22282566

**Authors:** Dominic Padova, Andreia Faria, J. Tilak Ratnanather, Raymond So, Stanley Zhu, Yuri Agrawal

**Affiliations:** Department of Otolaryngology – Head and Neck Surgery, Johns Hopkins University School of Medicine, Baltimore, MD, USA; Department of Radiology, Johns Hopkins University School of Medicine, Baltimore, MD, USA; Center for Imaging Science and Institute for Computational Medicine, Department of Biomedical Engineering, Johns Hopkins University, Baltimore, MD, USA; Department of Biomedical Engineering, Johns Hopkins University, Baltimore, MD, USA

**Keywords:** Vestibular, VEMP, VOR, MRI, Volume, Cortical, Cross-sectional

## Abstract

The vestibular system is emerging as a pre-eminent contributor to alterations in the structure and function of the central nervous system. Yet, whether age-related vestibular loss is related to volume loss in the cerebral cortical areas that receive vestibular input remains unknown. In this cross-sectional study of 117 healthy, older adults from the Baltimore Longitudinal Study of Aging, we examine the relationships between age-related vestibular functions and the gray matter volumes of the prefrontal cortex and its subregions and of the sensorimotor cortex—regions known to process vestibular information. To measure the functions of three vestibular organs, the saccule, utricle, and horizontal semi-circular canal vestibulo-ocular reflex (VOR), we performed cervical vestibular-evoked reflex (cVEMP), ocular VEMP (oVEMP), and video-head impulse tests, respectively. Log-linear multiple regression was used to investigate the relationships between average regional volume and vestibular function, adjusting for age, sex, and intracranial volume. We found that age-related changes in vestibular end-organ function differentially alter gray matter volumes in the prefrontal and sensorimotor cortices, with many findings persisting when considering left (or right) side only. Lower canal function had a degenerative effect on the volume of the prefrontal cortex concomitant with ongoing, age-related, global brain atrophy. Lower saccular function preserved the volume of the sensorimotor cortex against age-related, global brain atrophy and had no relationship with the prefrontal cortical volume. Whereas lower utricular function showed a degenerative effect on the volume of the middle frontal gyrus accompanying age-related, global brain atrophy, it showed a protective effect on the volume of the pole of the superior frontal gyrus and showed no relationship with sensorimotor cortical volume. Together, these findings suggest that vestibular function may play a role in the resilience to or acceleration of global age effects on regional brain atrophy. Moreover, these findings enhance the understanding of the role of age-related vestibular function in the structural alterations of the cerebral cortex.

**Key Points:**

- Age-related vestibular function is significantly associated with gray matter volumes in the prefrontal and sensorimotor cortices in adults.
- Lower canal function showed a degenerative effect on the volume of the prefrontal cortex in addition to ongoing age-related brain atrophy. Lower saccular function had a protective effect against age-related atrophy on sensorimotor cortical volume. Lower utricular function showed a degenerative effect on the relative volume of the middle frontal gyrus and a protective effect on the relative volume of the pole of the superior frontal gyrus. Canal and utricular function were not associated with the relative volumes of the sensorimotor cortex, and saccular function was not associated with the relative volumes of the prefrontal cortex.
- Lower canal and utricular function may play an important role in the acceleration of age-related brain atrophy in the prefrontal cortex and in the middle frontal gyrus, respectively. Lower utricular function may play a role in the resilience to age-related atrophy in the pole of the superior frontal gyrus.

## 1. Introduction

The vestibular, or inner ear balance, system is comprised of five organs: three semi-circular canals which detect rotational accelerations of the head in three dimensions, and two otolith organs, the utricle and saccule, which detect linear head accelerations in three dimensions and the orientation of the head with respect to gravity (Smith, 2019). Vestibular information is transmitted from the inner ear via the 8th nerve to the vestibular nuclear complex in the brainstem and to the cerebellum. Then, the vestibular nuclei pass vestibular input to descending pathways that facilitate postural and ocular reflexes which respectively help to maintain stable balance and gaze control during movement (Cullen, 2019). Additionally, vestibular information from the brainstem is sent through ascending pathways to the thalamus—primarily to the ventroposterior, ventrolateral, and anterior dorsal nuclei. From the thalamus, vestibular information is sent along three pathways in parallel: to the hippocampus, to the somatosensory and motor cortices, and to the prefrontal cortex (De Waele, Baudonnière, Lepecq, Huy, & Vidal, 2001; Hitier, Besnard, & Smith, 2014; Lopez, C. & Blanke, 2011). Notably, evidence is mounting that the vestibular system contributes to visuospatial cognitive function (Bigelow & Agrawal, 2015; Cullen, 2014; Smith, 2019; Smith & Zheng, 2013; Yoder & Taube, 2014), attention, executive function, and memory function (Bigelow & Agrawal, 2015). Vestibular structure (Lopez, Ivan, Honrubia, & Baloh, 1997; Richter, 1980) and function (Agrawal, Carey, Della Santina, Schubert, & Minor, 2009; Paige, 1992; Zalewski, 2015) decline with normal aging, and vestibular loss is associated with cognitive declines in older adults (Anson et al., 2019; Bigelow & Agrawal, 2015; Bigelow et al., 2015; Semenov, Bigelow, Xue, du Lac, & Agrawal, 2016; Xie et al., 2017). However, the exact anatomical pathways by which vestibular loss may impact cognition in older adults are largely unknown.

Numerous clinical studies have shown a link between peripheral vestibular loss and functional and structural changes in brain regions involved in vestibular processing and compensation. Brandt et al. demonstrated that, in subjects with bilateral vestibulopathy (BVP), lack of vestibular input leads to a 17% reduction in manually measured bilateral hippocampal volume relative to controls that correlated with impaired spatial memory and navigation assessed by the virtual Morris water navigation task (vMWT) (Brandt et al., 2005). In contrast to manual volumetry of gray matter, voxel-based morphometry (VBM) is an automated approach used to measure voxel-wise differences in relative concentrations of gray matter between two groups of subjects. In a VBM study comparing patients with complete unilateral vestibular deafferentation to healthy controls, Hüfner et al. reported reductions in gray matter volume (GMV) following vestibular nerve section in the cerebellum, supramarginal gyrus on the same side as the lesion, postcentral and superior temporal gyri, and in motion-sensitive visual area MT/V5 (Hüfner et al., 2009). Using VBM to compare patients recovering from vestibular neuritis to age-matched controls, Helmchen et al. found GMV increases in the insula, superior temporal gyrus, and retroinsular region which correlated with improving clinical vestibular assessments, GMV increases in the inferior parietal lobe, midcingulate cortex, cerebellum, and MT/V5, and a decrease in GMV in the midline pontomedullary junction (Helmchen et al., 2009). zu Eulenberg et al. found that patients with an acute unilateral vestibular deficit resulting from vestibular neuritis showed GMV reduction in the left posterior hippocampus, the right superior temporal gyrus, and the right superior frontal gyrus regardless of the side of vestibular impairment, as well as increases in the medial vestibular nuclei and MT/V5 bilaterally and right-side gracile nucleus (zu Eulenburg, Stoeter, & Dieterich, 2010). In a longitudinal VBM study of patients recovering from unilateral vestibular neuritis, Hong et al. observed increases in GMV in the right inferior frontal gyrus, right insula, right superior temporal sulcus, right lingual gyrus, left middle and inferior occipital gyrus, bilateral hippocampus, bilateral parahippocampal gyri, left caudate nucleus, bilateral cerebellum, and right flocculus and decreases in GMV in the right superior medial gyrus, right middle orbital gyrus, cerebellar vermis, and right cerebellar hemisphere (Hong, Kim, Kim, & Lee, 2014). Göttlich et al. used VBM and found GMV decreases in the bilateral hippocampal subfield CA3 with increasing vestibulopathy-related impairment in patients with incomplete BVP compared to healthy age- and gender-matched controls (Göttlich et al., 2016). In a VBM study of patients with BVP and age- and gender-matched controls, Kremmyda et al. found that vestibular loss due to incomplete BVP leads to GMV decreases bilaterally in the middle hippocampus and posterior parahippocampus, to deficits in objective and subjective evaluations of spatial memory and navigation performance using the vMWT and questionnaires, and to higher spatial anxiety (Kremmyda et al., 2016). Wurthmann et al. demonstrated using VBM that patients with persistent postural perceptual dizziness exhibited reductions in GMV in the left superior temporal gyrus, left MT/V5, bilateral middle temporal gyrus, bilateral cerebellum, left posterior hippocampus, right precentral gyrus, left anterior cingulate cortex, left caudate nucleus, and left dorsolateral prefrontal cortex (Wurthmann et al., 2017).

Additionally, several studies have demonstrated relationships between age-related, subclinical loss of vestibular end-organ functions and brain structure. Age-related loss of saccular function was associated with GMV loss in the hippocampus (Jacob et al., 2020; Kamil, Jacob, Ratnanather, Resnick, & Agrawal, 2018) and the left entorhinal cortex (Jacob et al., 2020) in cross-sectional studies of study of healthy, older adults. Jacob et al. also found surface contraction of the ventral lateral nucleus and reticular nucleus of the right thalamus, of ventral and ventrolateral regions of the caudate and putamen, respectively, in the left-side basal ganglia (Jacob et al., 2020). Furthermore, Jacob et al. found surface expansions of the CA1 and CA2 subfields of the left hippocampus, of the basolateral and basomedial nuclei in the left amygdala, of the left entorhinal cortex, and of the left entorhinal-transentorhinal cortical complex (Jacob et al., 2020). In a longitudinal study of adults, Padova et al. found that poorer saccular function was associated with GMV loss of the thalamus and basal ganglia, and horizontal semicircular canal sensory loss was associated with GMV loss in the left hippocampus (Padova, Ratnanather, Xue, Resnick, & Agrawal, 2020). Though the understanding of how peripheral vestibular end-organ functions lead to structural changes in the brain has improved, a key question yet to be answered is: how does age-related vestibular function affect cortical brain structures receiving vestibular inputs?

This cross-sectional study investigates whether age-related vestibular end-organ functions predict gray matter volumes of several cortical structures known to process vestibular information: the prefrontal cortex, comprised of the superior, middle, and inferior frontal gyri, and the precentral and postcentral gyri of the sensorimotor cortex. Multiple log-linear regression adjusted for age, intracranial volume, and sex is used to investigate the relationships between saccular, utricular, and horizontal semi-circular canal functions and MRI-based volumes of 117 of healthy, older participants aged 60 years and older from the Baltimore Longitudinal Study of Aging between 2013 and 2015.

## 2. Materials and Methods

### 2.1. Data

The data is a subset from the Baltimore Longitudinal Study of Aging (BLSA) (Shock & Gerontology Research Center, 1984) involving 117 participants ≥ 60 years old who had MRI brain scans and vestibular testing in the same visit between 2013 and 2015. All participants gave written informed consent, and none had any history of psychiatric disorders or were diagnosed with a vestibular, ophthalmological, microvascular, or neurodegenerative disease. Hearing loss was measured as the speech-frequency pure tone average of air-conduction thresholds at 0.5, 1, 2, and 4 kHz from the better ear and was included as a confounding variable in the supplemental analysis.

### 2.2. Vestibular physiologic testing

Vestibular function testing included measurement of saccular function using the cervical vestibular-evoked myogenic potential (cVEMP) test, of utricular function using the ocular VEMP (oVEMP) test, and of horizontal semicircular canal function using video head-impulse test (vHIT). cVEMP and oVEMP signals were recorded using a commercial electromyographic system (software version 14.1, Carefusion Synergy, Dublin, OH) (Li, Zuniga, Nguyen, Carey, & Agrawal, 2014; Nguyen et al., 2010) and were amplified ×2500 and band-pass filtered for the 20-2000 Hz and 3-500 Hz frequency intervals, respectively. Electromyograms (EMG) were recorded by disposable, pre-gelled Ag/AgCl electrodes set with 40-inch safety lead wires from GN Otometrics (Schaumburg, IL).

#### 2.2.1. Cervical vestibular-evoked myogenic potential (cVEMP)

The cVEMP test measures the function of the saccule (and inferior vestibular nerve), following published procedures (Harun, Oh, Bigelow, Studenski, & Agrawal, 2016; Li et al., 2014; Li et al., 2015; Nguyen et al., 2010). Participants sat on a chair inclined at 30°above the horizontal plane. Trained examiners placed EMG electrodes bilaterally on the sternocleidomastoid and sternoclavicular junction. A ground electrode was positioned on the manubrium sterni. The examiners instructed participants to turn their heads to generate at least a 30 *μ*V background response prior to delivering sound stimuli. Auditory stimuli of 500 Hz and 125 dB were administered in bursts of 100 stimuli monoaurally through headphones (VIASYS Healthcare, Madison, WI). cVEMPs were recorded as short-latency EMGs of the inhibitory response of the ipsilateral sternocleidomastoid muscle. Corrected cVEMP amplitudes were calculated by removing nuisance background EMG activity collected 10 ms prior to the onset of the auditory stimulus. The presence or absence of a cVEMP response, respectively indicating normal or impaired saccular function, was defined per established amplitude and latency thresholds (Li et al., 2014; Nguyen et al., 2010). The higher corrected cVEMP amplitude (unitless) from the left and right sides was used as a continuous measure of saccular function. A difference of 0.5 in corrected cVEMP was considered clinically relevant, in accordance with the literature (Nguyen et al., 2010).

#### 2.2.2. Ocular vestibular-evoked myogenic potential (oVEMP)

The oVEMP test measures the function of the utricle (and superior vestibular nerve), following established procedures (Harun et al., 2016; Li et al., 2014; Li et al., 2015; Nguyen et al., 2010). Participants sat on a chair inclined at 30°above the horizontal plane. Trained examiners placed a noninverting electrode ≈3 mm below the eye centered below the pupil and an inverting electrode 2 cm below the noninverting electrode. A ground electrode was placed on the manubrium sterni. Before stimulation, participants were instructed to perform multiple 20°vertical saccades to ensure that symmetric signals are recorded from both eyes. Participants were asked to maintain an upgaze of 20°during oVEMP testing and recording. Vibration stimuli included head taps applied to the midline of the face at the hairline and ≈30% of the distance between the inion and nasion using a reflex hammer (Aesculap model ACO12C, Center Valley, PA). oVEMPs were recorded as short-latency EMGs of the excitation response of the contralateral external oblique muscle of the eye. The presence or absence of an oVEMP response, respectively indicating normal or impaired utricular function, was defined per established amplitude and latency thresholds (Li et al., 2014; Nguyen et al., 2010). The higher oVEMP amplitude (*μ*V) from the left and right sides was used as a continuous measure of utricular function. A difference of 5 *μV* in oVEMP was considered clinically relevant, in accordance with the literature (Nguyen et al., 2010).

#### 2.2.3. Video head impulse test (vHIT)

The vHIT measures the horizontal vestibular-ocular reflex (VOR) (Agrawal, Davalos-Bichara, Zuniga, & Carey, 2013; Agrawal et al., 2014; Harun et al., 2016), and was performed using the EyeSeeCam system (Interacoustics, Eden Prarie, MN) in the same plane as the right and left horizontal semicircular canals (Agrawal et al., 2014; Schneider et al., 2009). To position the horizontal canals in the plane of stimulation, trained examiners tilted the participant’s head downward 30°below the horizontal plane. Participants were asked to maintain their gaze on a wall target ≈1.5 m away. A trained examiner delivers rotations of 5-10°(≈150-250°per second) to the participant’s head. The head impulses are performed at least 10 times parallel to the ground toward the right and left, chosen randomly for unpredictability. The EyeSeeCam system quantified eye and head velocity. VOR gain was calculated as the unitless ratio of the eye velocity to the head velocity. A normal VOR gain is equal to 1.0, indicating equal eye and head velocities. Hypofunction of the semi-circular canals is indicated by a VOR gain <0.8 accompanied by refixation saccades (Agrawal et al., 2013; Li et al., 2015; Weber, MacDougall, Halmagyi, & Curthoys, 2009). The mean VOR gain from the left and right sides was used as a continuous variable. A difference of 0.1 in VOR gain was considered clinically relevant, in accordance with the literature (Harun et al., 2016).

### 2.3. Structural MRI acquisition, processing, and quality control

T1-weighted volumetric MRI scans were acquired in the sagittal plane using a 3T Philips Achieva scanner at the National Institute on Aging Clinical Research Unit. The sequence used was a T1-weighted image (WI) (magnetization prepared rapid acquisition with gradient echo (MPRAGE); repetition time (TR)=6.5 ms, echo time (TE)=3.1 ms, flip angle=8°, image matrix=256×256, 170 slices, voxel area=1.0×1.0 mm, 1.2 mm slice thickness, FOV=256×240 mm, sagittal acquisition). Scans were automatically segmented using MRICloud (https://www.mricloud.org/) and the T1 multi-atlas set “BIOCARD3T_297labels_10atlases_am_hi_erc_M2_252_V1”. A semi-automated quality control pipeline was used to automatically identify and manually exclude scans and segmentations of poor quality. The general quality of the scans and segmentations were evaluated by three trained raters (D.P., R.O., and S.Z.). One scan was removed due to motion artefacts. To identify possible regionally specific errors left- and right-side volumes for each region of interest (ROI) in MNI space were residualized with respect to intracranial volume (Pomponio et al., 2020) and converted to z-scores. Absolute z-score distances greater than the threshold of 3 standard deviations were flagged for manual verification. One hemispheric region of interest from three participants (left posterior superior frontal gyrus, right opercularis, and left orbitalis) were flagged and verified by visual inspection as having segmentation errors. These three segmentations were subsequently removed rather than fixed because there are no consistent landmarks to guide the manual correction of these ROIs. Volumes were calculated by the number of voxels in each ROI multiplied by the size of a voxel in *mm*^3^. Intracranial volume was comprised of bilateral cerebral volumes, cerebellum, brainstem, and cerebrospinal fluid. Our analysis focuses on the ROIs relevant to our hypothesis, according to the hierarchy defined in JHU-MNI-SS brain (Oishi et al., 2009) and shown in Figure 1 and Table 2A:

**Figure 1.**
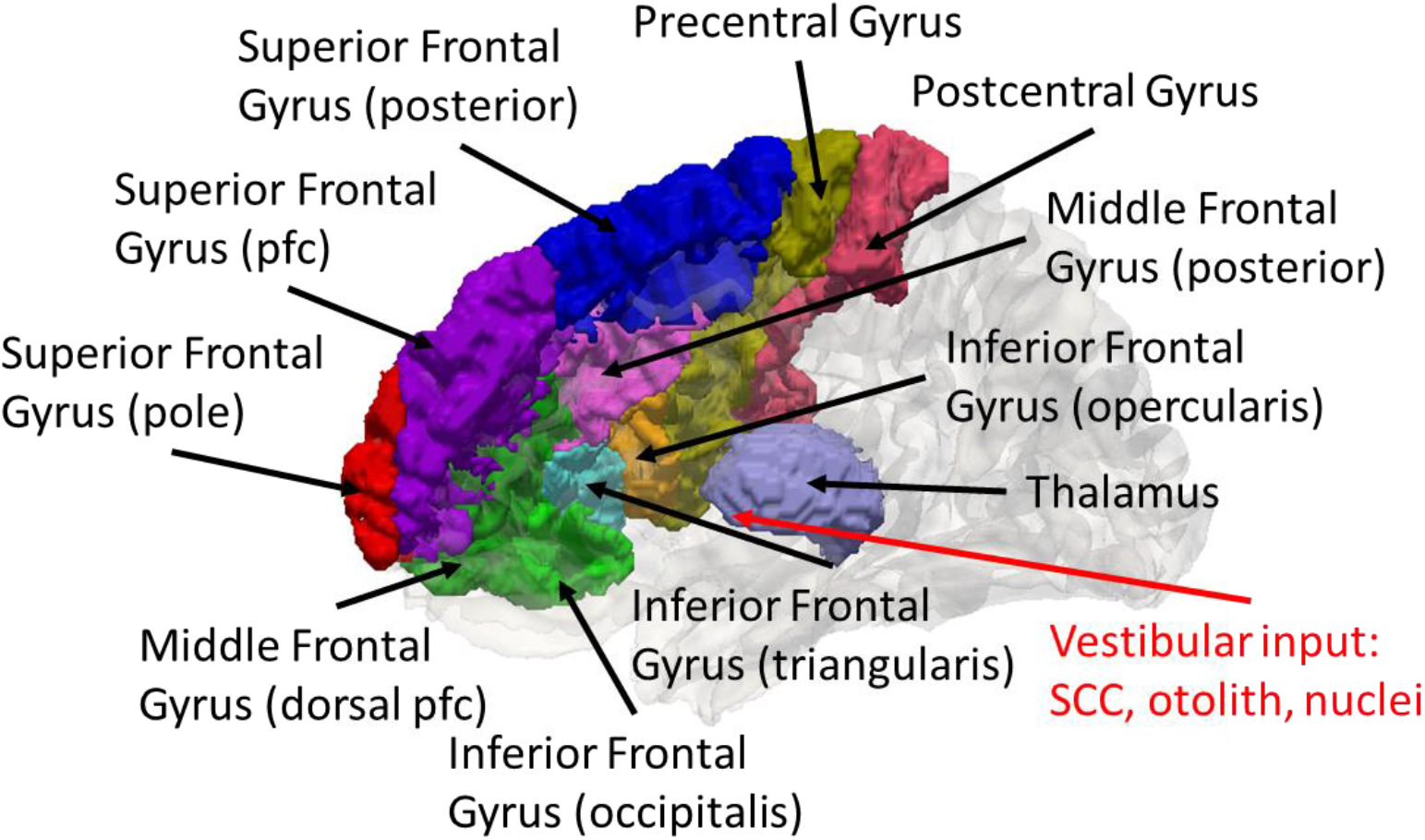
Cortical targets of the vestibular-thalamus-prefrontal cortical circuit. Vestibular information from the semicircular canals, otoliths, and nuclei is relayed through the thalamus to the sensorimotor cortex (pre and postcentral gyrus) and in turn to the prefrontal cortex (frontal gyrus). The red arrow points toward the ventral lateral nucleus, a putative subfield of the thalamus that receives vestibular input (Jacob et al., 2020). CAWorks (www.cis.jhu.edu/software/caworks) was used for visualization. Key: pfc: prefrontal cortex; SCC: semicircular canals.

1) Prefrontal cortex, composed of
1a) Superior frontal gyrus: defined as the sum of middle-superior part of the prefrontal cortex, frontal pole, and posterior pars of superior frontal
1b) Middle Frontal gyrus: defined by the sum of dorsal prefrontal cortex and posterior pars of middle frontal
1c) Inferior frontal gyrus: defined as the sum of pars opercularis, triangularis, and orbitalis of inferior frontal.
2) Sensorimotor cortex, composed of
2a) Precentral Gyrus
2b) Postcentral Gyrus

### 2.4. Statistical modeling

For participants with missing vestibular data, we carried over data from an adjacent prior or subsequent visit (Mongin et al., 2019). Whereas our original dataset had 58, 64, and 91 observations for cVEMP, oVEMP, and VOR, respectively, the imputed dataset had 95, 100, 107 observations for cVEMP, oVEMP, and VOR, respectively. Using this imputed dataset, multiple generalized linear regression adjusted for age, intracranial volume, and sex was used to investigate the relationship between regional volume and vestibular function. The null hypothesis in Eq. (1) predicts the natural logarithm of regional volume *voli*, for participant *i, i* = 1, …, *N*. The alternate hypothesis predicts the natural logarithm of regional volume using a vestibular variable, *vest*_*i*_ in Eq. (2), such as best corrected cVEMP, best oVEMP, and mean VOR gain as continuous independent variables,

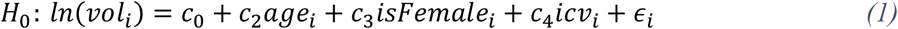

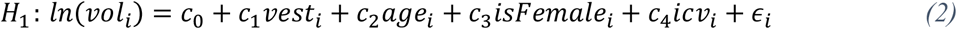

where *c*_0_ corresponds to the global average, *age*_*i,j*_ of subject *i, isFemale*_*i*_ is a binary indicator variable for the sex of subject *i* (1=female, 0=male), and *icν*_*i*_ denotes the intracranial volume of subject *i*. We assumed that the natural logarithm of regional volume depends linearly on age. We also assumed that the measurement noise *∈*_*i*_ is independently and identically distributed zero-mean Gaussian with unknown, common variance. The unknown effects {*c*_0_, *c*_1_, *c*_2_, *c*_3_, *c*_4_} were estimated via maximum likelihood. The vestibular effects on the linear scale were multiplied by their respective relevant clinical effect size (corrected cVEMP: 0.5, oVEMP: 5 *μV*, VOR: 0.1). In turn, the effects are interpreted as the relative regional volume change resulting from a 1-clinically-relevant-unit change in vestibular variable. All effects reported have been transformed according to *β*_*k*_ = 100(exp(*c*_*k*_) ™ 1), *k* = 0, …, 4. To determine whether the study sample is stable and that our individual results are not driven by outliers or extreme values, we performed a permutation likelihood ratio test (LRT) by permuting model residuals under the null hypothesis, *H*_0_. The permuted p-value, *p*_*perm*_, was calculated as the proportion of simulated LRTs, termed *LRT*_*sim*_, greater than the observed LRTs, termed *LRT*_*obs*_, (*n*_*perm*_ = 5000 simulations),

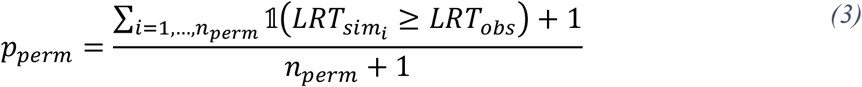

where 𝟙(*x*) is the indicator function that yields 1 when the argument *x* is true and 0 otherwise. We reject the null hypothesis if *p*_*perm*_ < 0.05. For models which rejected the null hypothesis, 95% confidence intervals were calculated by bootstrapping model residuals under the alternative hypothesis, *H*_1_, (10k simulations) to mitigate the effects of outliers. A Benjamini-Hotchberg procedure was used to control the false discovery rate (FDR) of the comparisons made in this study (Benjamini & Hochberg, 1995). FDR q-values indicate the expected proportion of rejected null hypotheses that are false. To avoid being overly conservative in our adjustment for multiple comparisons, we chose to adjust FDR for each side of the brain and for each level of granularity separately. For these exploratory analyses, we considered FDR thresholds of 0.05 and 0.10. All effects were considered significant at the *q* < 0.05 and *q* < 0.1 levels. These analyses were implemented in *RStudio* (RStudio Team, 2020). Bootstrapped confidence intervals were computed using the *np*.*boot* function of the nptest R package (Helwig, 2019).

## 3. Results

### 3.1. Study sample characteristics

Table 1 shows the characteristics for the study sample from the Baltimore Longitudinal Study of Aging.

**Table 1.** Characteristics of the study sample (n = 117). Key: PTA: four-frequency (0.5, 1, 2, 4 kHz) pure tone average from the better ear; N: the number of participants with a visit where both the characteristic and MRI data were available; %: 100(N/n) percent; SD: standard deviation.

| Characteristic | Mean (SD) | N (%) |
| --- | --- | --- |
| Age (years) | 77 (8.7) |  |
| Sex |  |  |
| Male |  | 79 (67.5) |
| Female |  | 38 (32.5) |
| Corrected cVEMP Amplitude | 1.2 (0.749) |  |
| oVEMP Amplitude ($\mu V$) | 13.6 (10.1) | |
| Mean VOR Gain | 0.997 (0.163) |  |
| Four Frequency PTA (dB) | 32 (14.9) |  |

### 3.2. Association between vestibular function and regional cortical volumes based in generalized linear regression

Figure 2A summarizes the relationships between the definitions of the anatomical regions of interest, where the child nodes are a part of the parent nodes, as illustrated in Figure 1. Figure 2B depicts the associations between vestibular functions and regional brain volumes for models which rejected the null hypothesis according to permutation testing. The permutation testing results show that our sample is stable: our individual results are not driven by outliers or extreme values. Because age and intracranial volume are included as covariates, the vestibular effects on regional volume may be interpreted as percent volume preservation in the face of global, age-related volume reductions for each 1-clinical-unit increase in the vestibular variable. This means that regions that exhibit a higher relative volume do not actually have a higher volume; rather, they have less age-related atrophy than the whole brain, meaning they are relatively spared. Figure 2C summarizes the raw p-values calculated from the observed (not permuted) data and corresponding FDR-corrected q-values for each vestibular-volume comparison for each side at each level of granularity for the prefrontal cortex and the sensorimotor cortex. A total of 42 regression models were assessed and 18 regression models rejected the null hypothesis at the 0.05 level according to permutation testing (See Figure 2B). To correct for multiple independent comparisons across regions of interest, we performed an additional analysis of the raw p-values estimated from the 42 observed data models: seven relationships were significant (i.e. remained significant after FDR-correction) at the 0.1 level (See Figure 2C). In this additional analysis, three relationships at the coarsest level of granularity were significant at the q < 0.05 level: poorer canal function and lower relative volumes of the bilateral (% change ≈ 1.12, q-value ≈ 0.04), left (% change ≈ 1.13, q-value ≈ 0.045), and right (% change ≈ 1.11, q-value ≈ 0.048) prefrontal cortex. At the coarse-fine granularity, the relationship between poorer canal function and smaller relative volume of the left superior frontal gyrus was significant at the 0.05 level (% change ≈ 1.55, q-value ≈ 0.046) and relationship between poorer utricular function and lower relative volume of the right middle frontal gyrus (% change ≈ 1.16, q-value ≈ 0.086) was significant at the 0.1 level. Two relationships at the finest granularity were significant at the 0.1 level: the poorer saccular function and higher relative volume of the left postcentral gyrus (% change ≈ -2.22, q-value ≈ 0.086) and poorer utricular function and higher relative volume of the pole of the left superior frontal gyrus model (% change ≈ -2.01, q-value ≈ 0.084). Supplementary Table S1 details the numerical values of the vestibular relationships with relative regional volume, including the numerical values corresponding to Figures 2B and 2C. Figure 3 shows the spatial arrangement of the average estimated vestibular effects for models which remained significant after FDR-control at the 0.05 and 0.01 levels.

**Figure 2.**
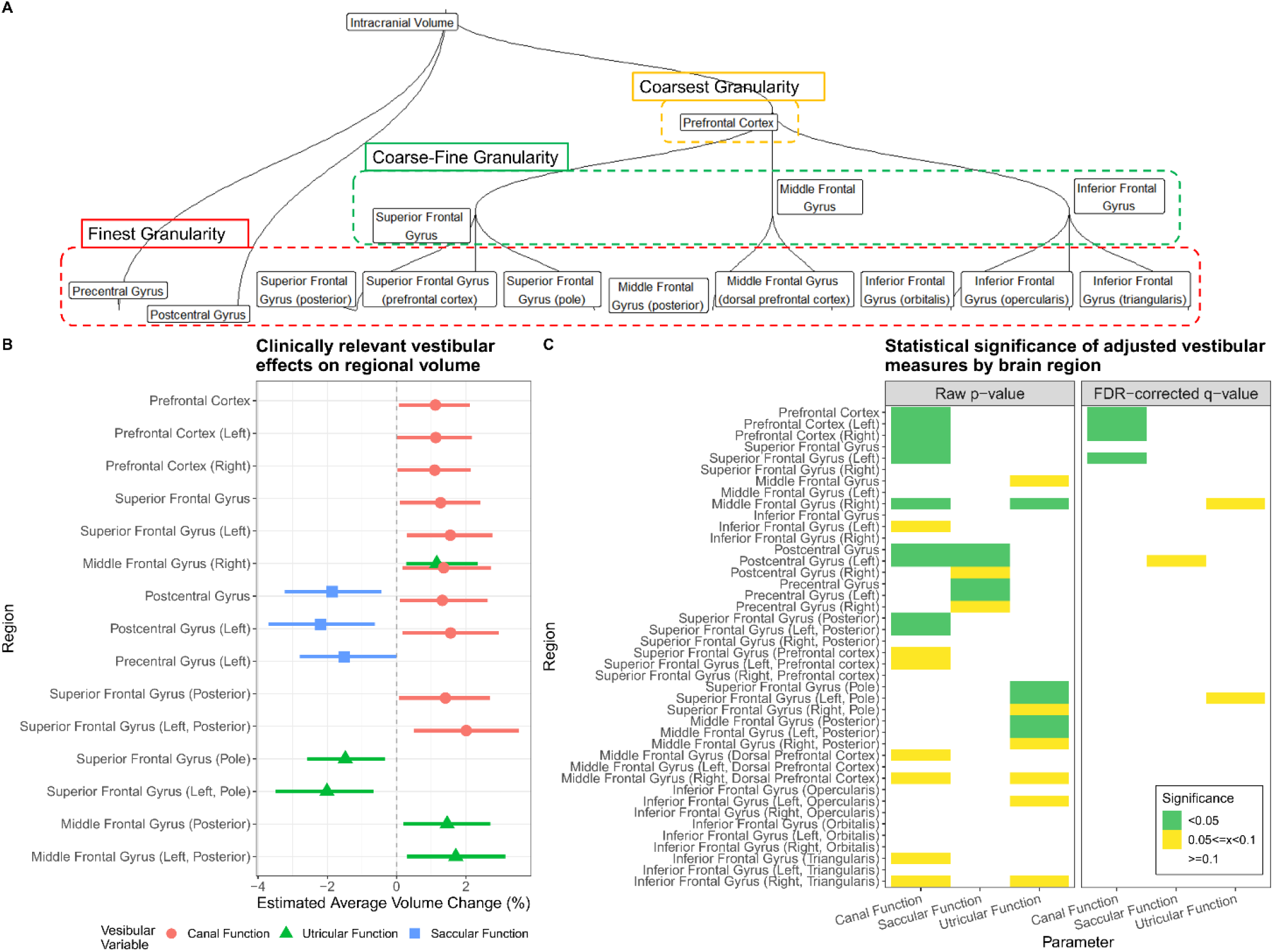
(A) A dendrogram showing the hierarchical relationships between the labels in which the brains were segmented. (B) A dot and whisker plot of the vestibular effects (estimated average volume preservation in percent) on regional gray matter volumes with respect to a 1-clinically-relevant-unit increase in vestibular function for models which rejected the null hypothesis according to permutation testing. The point-shapes correspond to the average effect and the extent of the whiskers corresponds to the 95% confidence intervals. Notably, regions that exhibit a higher relative volume do not actually have a higher volume; rather, they have less age-related atrophy than the whole brain—they are relatively spared. (C) Heat maps of the raw p-values (left) calculated from the observed data models and the FDR-corrected q-values (right) for all granularities of structure models by region. FDR-correction was performed independently for each side (bilateral-mean, left, right) and level of granularity. All models were adjusted for age, sex, and intracranial volume (n=117).

**Figure 3.**
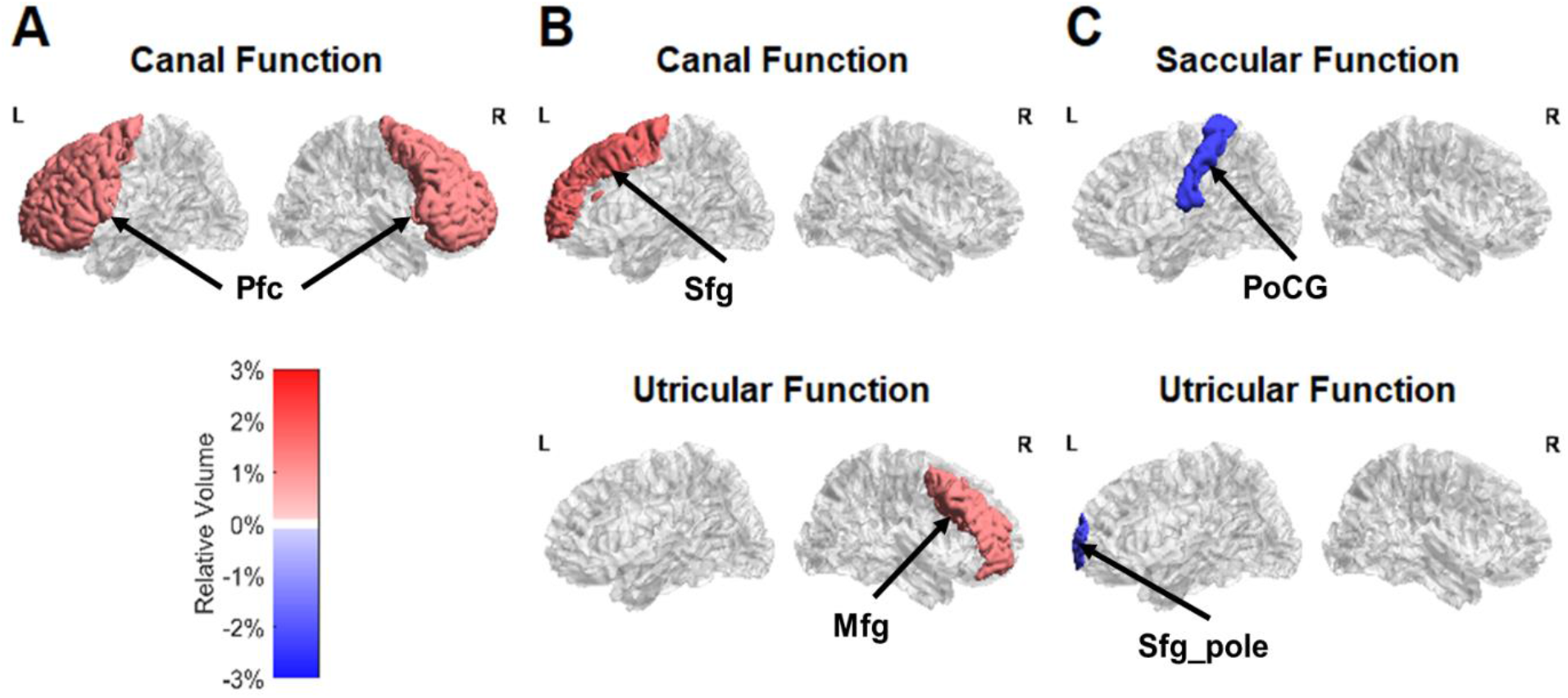
Spatial distribution of average vestibular effects on regional gray matter volumes at three levels of granularity with respect to a 1-clinically-relevant-unit increase in vestibular function for models which survived FDR-control at the 0.05 and 0.1 levels. (A) coarse granularity: from the top row downwards, the prefrontal cortex (Pfc) and color bar. (B) coarse-fine granularity: left superior frontal gyrus (Sfg) (top row) and right middle frontal gyrus (Mfg) (middle row). (C) fine granularity: left postcentral gyrus (PoCG) (top row), left pole of the superior frontal gyrus (Sfg_pole) (bottom row). Whereas red color indicates percent volume expansion in the face of age-related global brain size reduction, blue color indicates relative volume reduction. Notably, regions that exhibit a higher relative volume do not actually have a higher volume; rather, they have less age-related atrophy than the whole brain—they are relatively spared. The L and R labels indicate the lateral views of the left and right hemispheres, respectively. This figure was visualized with the BrainNet Viewer [9] (http://www.nitrc.org/projects/bnv/).

To examine whether the addition of hearing function can account for the potential relationships between vestibular function and cortical volumes, we performed our analysis pipeline additionally adjusting for hearing function. The addition of hearing function to the models reduced the sample size from 117 to 115 participants and resulted in the same significant associations previously observed with similar effect sizes (See Supplementary Table S2).

## 4. Discussion

In this study, we investigated the relationship between vestibular function and cortical gray matter volume (GMV) in older adults. We examined key cortical regions involved in vestibular processing: the prefrontal cortex, made up of the superior, middle, and inferior frontal gyri, the precentral gyrus in the motor cortex, and the postcentral gyrus in the somatosensory cortex (De Waele et al., 2001; Hitier et al., 2014; Lopez & Blanke, 2011). We found significant relationships between vestibular function and the volumes of these cortical areas. To our knowledge, these findings provide some of the first demonstrations that age-related loss of vestibular end-organ functions in older adults is associated with GMV changes in several prefrontal and sensorimotor cortical regions that receive vestibular inputs. Notably, we found in this cohort of healthy older adults that vestibular impairment (both canal and otolith) was associated with reduced GMV in the prefrontal cortex, and specifically the superior and middle frontal gyri. These findings are compatible with clinical studies that demonstrated that patients with vestibular impairment had reductions in GMV in the right superior frontal gyri (Hong et al., 2014) and in the left dorsolateral prefrontal cortex compared to controls (Wurthmann et al., 2017). Furthermore, neuroimaging during vestibular stimulation studies in humans have shown activations in regions of the prefrontal and sensorimotor cortices (Lopez & Blanke, 2011) in which we observed GMV alterations.

### 4.1. Cortical areas of vestibular-modified gray matter volume alterations

#### 4.1.1. Prefrontal cortex

The prefrontal cortex showed a lower GMV with poorer canal function. The prefrontal cortex is thought to be involved in executive function (Yuan & Raz, 2014), which encompasses planning, decision-making, working memory, inhibitory control, and abstract rule-learning. Peripheral vestibular information is transmitted to the brainstem which sends this information to the prefrontal cortex via thalamic-limbic-striatal-frontal circuits. Although the exact functions of the circuits are poorly defined, vestibular function has been shown to be associated with working memory in older adults (Bigelow et al., 2015) and to mental number ordering and manipulation ability (Grabherr, Cuffel, Guyot, & Mast, 2011; Moser, Vibert, Caversaccio, & Mast, 2017; Preuss, Mast, & Hasler, 2014).

We observed a lower GMV of the superior frontal gyrus with poorer canal function, as well as lower GMV of the middle frontal gyrus with poorer utricular function. Each of these impacted areas serves potentially many important roles in a person’s ability to interact with the world. Toward the posterior end, the superior frontal gyrus contains portions of the supplementary motor area and of the frontal eye field and is responsible in part for initiating complex movements. Owing to its circuitry and to containing portions of the premotor cortex, the middle frontal gyrus engages in aspects of motor control and planning. Interestingly, looking at a finer level of granularity, the anterior pole of the superior frontal gyrus showed a higher GMV with poorer utricular function. This more focal association was attenuated when considering the whole prefrontal cortex as well as the whole superior frontal gyrus, possibly due to relationships in the opposite direction involving the rest of those structures. Taken together, these results suggest that vestibular loss is related to relative atrophy of prefrontal cortical structures that subserve executive function and motor planning.

#### 4.1.2. Somatosensory and motor cortices

We observed an association between vestibular function and GMV of somatosensory cortex at the finest level of granularity. Specifically, the left postcentral gyrus showed a higher GMV with poorer saccular function. We observed no relationships after FDR-control between GMV in the precentral gyrus and vestibular function. The postcentral gyrus, a part of the somatosensory cortex within the parietal cortex, is topographically organized into maps of body parts (e.g., head, trunk) and receives projections from the thalamus which carry vestibular information. The postcentral gyrus is responsible for self-motion perception which contributes to motor planning (Anson, E., Pineault, Bair, Studenski, & Agrawal, 2019; Fasold et al., 2002; Lopez, C., Blanke, & Mast, 2012; Roditi & Crane, 2012; Stiles & Smith, 2015).

One explanation of this result could be that loss of saccular inputs results in reweighting by multisensory (e.g., visual, auditory, autonomic, proprioceptive) regions, like the vestibular nuclei, cerebellum, hypothalamus, and thalamus (Ferrè & Haggard, 2020; Saman, Arshad, Dutia, & Rea, 2020), before sending these reweighted inputs back to the postcentral gyrus. Such saccular modulation could suppress vestibular-mediated structural changes in the postcentral gyrus. Sensory substitution as a compensatory mechanism has been suggested to explain GMV increases (Helmchen et al., 2009; Hong et al., 2014). Additionally, the results indicating vestibular effects in opposing directions may be a product of different emphasis on the afferent and efferent projections that transmit vestibular information. For instance, the age-related loss of vestibular function associated with lower relative GMV may suggest a dominance of afferent connections receiving peripheral vestibular inputs, highlighting bottom-up processing. On the other hand, the age-related loss of vestibular function associated with higher relative GMV may suggest a dominance of efferent projections which modify remaining peripheral vestibular inputs that have declined due to aging. However, teasing out these bottom-up and top-down processes is a challenge that would require addressing the structural effects of compensatory and sensory substitution processes in brain regions that process vestibular and extra-vestibular information. Overall, these findings suggest differing roles of age-related vestibular end-organ functions in the morphological alterations in the cortex.

##### Interpretations of gray matter alterations

We note that the GMV changes can be interpreted as regional volume preservation effects. This connotation arises because the age and intracranial volume regressors account for linear slope trends in regional volume due to aging and global brain volume differences. This means that regions that exhibit a higher relative volume do not actually have a higher volume; rather, they have less age-related atrophy than the whole brain. Further interpretation of the nature of the effects is limited by the fact that the exact neural substrates underlying these GMV changes remains obscure. To uncover whether the vestibular effects reflect changes in neuronal density, supplemental histological and physiological studies would be needed. Moreover, the magnitudes of structural alterations must be understood with respect to a clinically relevant magnitude of vestibular function. In this study, clinically meaningful changes in saccular, utricular, and canal function measures are 0.5, 5*μV*, and 0.1, respectively. By multiplying the vestibular effects on the linear scale by their clinically relevant changes in value, each vestibular effect can be understood as the preservation or atrophy of GMV relative to ongoing age-related degeneration associated with a 1-clinical-unit change in vestibular function, or alternatively resilience to or acceleration of global age effects. Taken together, our findings suggest that lower vestibular function is primarily associated with accelerated atrophy overall in the prefrontal cortex and with resilience to regional brain atrophy in the postcentral gyrus of the sensorimotor cortex.

### 4.2. Strengths and limitations

We report several strengths of this study. One such strength is that the relationships examined were hypotheses-driven based on prior work, such as functional neuroimaging during vestibular stimulation and clinical studies in humans. A second strength is that our quality control pipeline involved manual inspections of the data at each step of processing. Third, our statistical testing pipeline accounts for multiple dependent and independent comparisons using permutation testing and FDR correction, respectively, as well as for outliers using bootstrapping. Still, there are limitations of this study. Because volumes are coarse measures of structure, it is possible that we did not detect true effects if they were nonuniformly distributed across or within the region of interest. Cortical thickness and surface shape may provide better measures of variation in the structure of the region of interest. This sensitivity becomes especially advantageous if the region of interest is convoluted and difficult to segment. Additionally, the reproducibility of results is a challenge at high cortical parcel granularity, as anatomical definitions can vary between atlases and experts. Another limitation is that we did not examine the potential role of the cerebellum, the brainstem, the hypothalamus, or the thalamus in modulating the effects of age-related vestibular loss in the cortex. However, robust measures of cerebellar and brainstem structures are being developed. Additionally, our findings may not generalize to the broader and younger population due to the age range used in this study and the propensity of BLSA participants to have higher levels of education and socioeconomic status than typical adults.

### 4.3. Future work

While this cross-sectional study helps to clarify the relationships between vestibular function and cortical regions of interest, subsequent longitudinal studies will be needed to elucidate how vestibular function may over time impact the structure of brain regions that receive vestibular input: the limbic system, temporo-parietal junction, and frontal cortex. Future work will also incorporate sensitive measures of local structure change, like cortical thickness and shape, which complement gross volume measures, as well as measures of microstructural change gleaned from diffusion MRI. Structural equation modeling of longitudinal data can be used to investigate causal hypotheses between vestibular loss and structural changes, simultaneously accounting for possible confounding by hearing and vision loss. This framework can also help to test hypotheses that intervening brain structures such as the brainstem, the hypothalamus, the cerebellum, and thalamus modify vestibular effects on downstream structures. Additionally, changepoint analysis can identify non-linearities in the trends of structural changes. The collection of changepoints for each structure measure can be temporally ordered to explain the sequence of structural changes in relation to each other. Ultimately, these studies together can reveal the sequence and causal direction of changes in the vestibular network.

## 5. Conclusion

This study found associations between age-related vestibular end-organ functions and regional GMVs of the prefrontal and somatosensory cortices in adults. This work furthers the understanding of the role of the vestibular system in structural changes in cortical regions that receive peripheral vestibular input and are affected in clinical vestibular disorders. Moreover, these vestibular-modified structural changes may provide anatomical links between vestibular function and cognition. Future work will need to determine the temporal and spatial flow of structural alterations in the central nervous system related to peripheral vestibular function.

## Data Availability

Data from the BLSA are available on request from the BLSA website (blsa.nih.gov). All requests are reviewed by the BLSA Data Sharing Proposal Review Committee and are also subject to approval from the NIH institutional review board.

https://blsa.nih.gov/

## Acknowledgements

This work was supported in part by the National Institute on Aging [grant number R01 AG057667], National Institute on Deafness and Other Communication Disorders [grant number R03 DC015583], and National Institutes of Health [grant number P41-EB031771] and by the Intramural Research Program, National Institute on Aging, National Institutes of health.

