## Supplemental Tables 1 and 2 for "Vestibular Function Predicts Prefrontal and Sensorimotor Cortical Gray Matter Volumes in a Cross-Sectional Study of Healthy, Older Adults"

**Supplemental Information**

**Table S1.** A summary of vestibular effects on relative regional volume for vestibular models that rejected the null hypothesis, each adjusted for age, sex, intracranial volume (n = 117). The vestibular estimate is the average value given in percent for a 1-clinically-relevant-unit increase in vestibular value, the 95% CI interval is given as (low, high) in units of percent, p-values are left uncorrected for multiple independent comparisons, and FDR-corrected q-values were obtained independently for each level of granularity and for each side (bilateral-mean, left, right). Notably, regions that exhibit a higher relative volume do not actually have a higher volume; rather, they have less age-related atrophy than the whole brain. o p,q < 0.1, * p,q < 0.05, ** p,q < 0.01, *** p,q < 0.001, **** p,q < 0.0001.

| Region | Vestibular Variable | Estimate | 95% CI | Raw p-value | q-value |
| --- | --- | --- | --- | --- | --- |
| Prefrontal Cortex | Canal Function | 1.12 | (0.0616, 2.11) | 0.0398 * | 0.0398 * |
| Prefrontal Cortex (Left) | Canal Function | 1.13 | (0.0204, 2.18) | 0.0452 * | 0.0452 * |
| Prefrontal Cortex (Right) | Canal Function | 1.1 | (0.0308, 2.14) | 0.0484 * | 0.0484 * |
| Superior Frontal Gyrus | Canal Function | 1.27 | (0.0874, 2.42) | 0.0375 * | 0.113 |
| Superior Frontal Gyrus (Left) | Canal Function | 1.55 | (0.295, 2.76) | 0.0154 * | 0.0462 * |
| Middle Frontal Gyrus (Right) | Utricular Function | 1.16 | (0.279, 2.35) | 0.0287 * | 0.0861 o |
| Middle Frontal Gyrus (Right) | Canal Function | 1.36 | (0.172, 2.73) | 0.0406 * | 0.122 |
| Postcentral Gyrus | Saccular Function | -1.86 | (-3.23, -0.433) | 0.014 * | 0.14 |
| Postcentral Gyrus | Canal Function | 1.32 | (0.0889, 2.63) | 0.0481 * | 0.168 |
| Postcentral Gyrus (Left) | Saccular Function | -2.2 | (-3.7, -0.625) | 0.00876 ** | 0.086 o |
| Postcentral Gyrus (Left) | Canal Function | 1.56 | (0.175, 2.95) | 0.0334 * | 0.17 |
| Precentral Gyrus (Left) | Saccular Function | -1.51 | (-2.8, -0.00716) | 0.0436 * | 0.218 |
| Superior Frontal Gyrus (Posterior) | Canal Function | 1.41 | (0.07, 2.7) | 0.0463 * | 0.168 |
| Superior Frontal Gyrus (Left, Posterior) | Canal Function | 2.01 | (0.489, 3.53) | 0.0114 * | 0.114 |
| Superior Frontal Gyrus (Pole) | Utricular Function | -1.48 | (-2.58, -0.33) | 0.0157 * | 0.134 |
| Superior Frontal Gyrus (Left, Pole) | Utricular Function | -2.01 | (-3.5, -0.668) | 0.00838 ** | 0.0838 o |
| Middle Frontal Gyrus (Posterior) | Utricular Function | 1.46 | (0.195, 2.71) | 0.0268 * | 0.134 |
| Middle Frontal Gyrus (Left, Posterior) | Utricular Function | 1.71 | (0.303, 3.14) | 0.0234 * | 0.117 |

**Table S2.** Vestibular and hearing predictors of relative regional volume (mm^3^) controlling for age, sex, intracranial volume, and the four-frequency pure tone average (PTA) (n = 115) with mean ± standard error (uncorrected p-value). The effects are reported as estimated average percent difference in relative volume corresponding to a 1-unit increase in the hearing parameter or a 1-clinically-relevant-unit increase in the vestibular parameter. Notably, regions that exhibit a higher relative volume do not actually have a higher volume; rather, they have less age-related atrophy than the whole brain. The speech-frequency PTA of air-conduction thresholds at 0.5, 1, 2, and 4 kHz from the better ear was used. *p < 0.05, **p < 0.01.

| Region | Vestibular Variable | Parameter | Effect of Parameter on Region |
| --- | --- | --- | --- |
| Prefrontal Cortex | Canal Function | Canal Function | 1.23 ± 0.562 (p=0.0318) * |
| Prefrontal Cortex | Canal Function | PTA | 0.00478 ± 0.0698 (p=0.946) |
| Prefrontal Cortex (Left) | Canal Function | Canal Function | 1.22 ± 0.583 (p=0.0403) * |
| Prefrontal Cortex (Left) | Canal Function | PTA | 0.0171 ± 0.0719 (p=0.813) |
| Prefrontal Cortex (Right) | Canal Function | Canal Function | 1.23 ± 0.575 (p=0.0358) * |
| Prefrontal Cortex (Right) | Canal Function | PTA | -0.00879 ± 0.0716 (p=0.903) |
| Superior Frontal Gyrus | Canal Function | Canal Function | 1.44 ± 0.631 (p=0.025) * |
| Superior Frontal Gyrus | Canal Function | PTA | 0.03 ± 0.0782 (p=0.702) |
| Superior Frontal Gyrus (Left) | Canal Function | Canal Function | 1.74 ± 0.658 (p=0.00993) ** |
| Superior Frontal Gyrus (Left) | Canal Function | PTA | 0.055 ± 0.0814 (p=0.501) |
| Middle Frontal Gyrus (Right) | Canal Function | Canal Function | 1.42 ± 0.689 (p=0.042) * |
| Middle Frontal Gyrus (Right) | Canal Function | PTA | -0.00548 ± 0.0863 (p=0.949) |
| Inferior Frontal Gyrus (Left) | Canal Function | Canal Function | 0.0205 ± 0.0896 (p=0.819) * |
| Inferior Frontal Gyrus (Left) | Canal Function | PTA | 1.78 ± 0.781 (p=0.0257) |
| Postcentral Gyrus | Saccular Function | Saccular Function | 0.0942 ± 0.094 (p=0.319) * |
| Postcentral Gyrus | Saccular Function | PTA | 0.0865 ± 0.0936 (p=0.358) |
| Postcentral Gyrus (Left) | Saccular Function | Saccular Function | -1.82 ± 0.827 (p=0.0282) * |
| Postcentral Gyrus (Left) | Saccular Function | PTA | 0.0663 ± 0.103 (p=0.521) |
| Precentral Gyrus (Left) | Saccular Function | Saccular Function | -2.08 ± 0.919 (p=0.024) * |
| Precentral Gyrus (Left) | Saccular Function | PTA | 0.0404 ± 0.114 (p=0.725) |
| Superior Frontal Gyrus (Posterior) | Canal Function | Canal Function | -1.66 ± 0.815 (p=0.0416) * |
| Superior Frontal Gyrus (Posterior) | Canal Function | PTA | 0.121 ± 0.102 (p=0.24) |
| Superior Frontal Gyrus (Left, Posterior) | Canal Function | Canal Function | 1.72 ± 0.729 (p=0.0211) ** |
| Superior Frontal Gyrus (Left, Posterior) | Canal Function | PTA | 0.0387 ± 0.0903 (p=0.669) |
| Superior Frontal Gyrus (Pole) | Utricular Function | Utricular Function | 2.32 ± 0.814 (p=0.00574) * |
| Superior Frontal Gyrus (Pole) | Utricular Function | PTA | 0.0807 ± 0.1 (p=0.424) |
| Superior Frontal Gyrus (Left, Pole) | Utricular Function | Utricular Function | -1.63 ± 0.676 (p=0.0164) ** |
| Superior Frontal Gyrus (Left, Pole) | Utricular Function | PTA | 0.00919 ± 0.0886 (p=0.918) |
| Middle Frontal Gyrus (Posterior) | Utricular Function | Utricular Function | -2.27 ± 0.842 (p=0.00738) * |
| Middle Frontal Gyrus (Posterior) | Utricular Function | PTA | -0.0144 ± 0.108 (p=0.895) |
| Middle Frontal Gyrus (Left, Posterior) | Utricular Function | Utricular Function | -0.0162 ± 0.0961 (p=0.866) * |
| Middle Frontal Gyrus (Left, Posterior) | Utricular Function | PTA | 1.91 ± 0.726 (p=0.0105) |
| Inferior Frontal Gyrus (Triangularis) | Canal Function | Canal Function | -0.0671 ± 0.119 (p=0.573) * |
| Inferior Frontal Gyrus (Triangularis) | Canal Function | PTA | 10.7 ± 5.69 (p=0.0431) |
| Inferior Frontal Gyrus (Right, Triangularis) | Utricular Function | Utricular Function | 0.154 ± 0.121 (p=0.206) * |
| Inferior Frontal Gyrus (Right, Triangularis) | Utricular Function | PTA | 15.4 ± 7.07 (p=0.0161) |
